# Transcriptomic signature on Hantavirus Cardiopulmonary Syndrome patients, reveals an increased interferon response as a hallmark of critically ill patients

**DOI:** 10.1101/2021.07.28.21260540

**Authors:** Grazielle E. Ribeiro, Eduardo Duran-Jara, Ruth Perez, Analia Cuiza, Luis E. Leon, Constanza Martínez-Valdebenito, Nicole Le Corre, Marcela Ferres, Leonila Ferreira, Maria Luisa Rioseco, Jorge Gavilán, Francisco Arancibia, Jerónimo Graf, Rene Lopez, Jose Luis Perez, Mario Calvo, Gregory J Mertz, Pablo A.Vial, Cecilia Vial, the hantavirus study group

**Affiliations:** Instituto de Ciencias e Innovación en Medicina, Facultad de Medicina, Clínica Alemana Universidad del Desarrollo, Santiago, Chile; Instituto de Ciencias Biomédicas, Facultad de Ciencias de la Salud, Universidad Autónoma de Chile, Santiago, Chile; Departamento de Enfermedades Infecciosas e Inmunología Pediátricas, Pontificia Universidad Católica de Chile, Santiago, Chile; Hospital Dr. Guillermo Grant Benavente Concepción, Concepción, Chile; Hospital Puerto Montt Dr. Eduardo Schütz Schroeder, Puerto Montt, Chile; Instituto Nacional del Tórax, Santiago, Chile; Departamento de Paciente Crítico, Clínica Alemana de Santiago, Santiago, Chile; Hospital Base San José de Osorno, Osorno, Chile; Instituto de Medicina, Universidad Austral de Chile - Hospital Base Valdivia, Valdivia, Chile; Department of Internal Medicine, University of New Mexico, Albuquerque, USA; Departamento Pediatría Clínica Alemana de Santiago, Santiago, Chile

## Abstract

New World hantaviruses are important human pathogens that can cause a severe zoonotic disease called hantavirus cardiopulmonary syndrome (HCPS). HCPS patients can progress quickly to a severe condition with respiratory failure and cardiogenic shock that can be fatal in 30% of the cases. The role of the host’s immune responses in this progression towards HCPS remains elusive. In this study, 12 patients hospitalized with severe HCPS were analyzed using a transcriptome approach combined with clinical laboratory data to gain a better insight into factors associated with a severe clinical course. Patients were further classified in two levels of severity, a first group that required mechanical ventilation and vasoactive drugs (VM+VD) and a second group that also needed ECMO or died (ECMO/Fatal). Their transcriptional profile was compared during acute (early and late) and convalescent phases. Our results showed that overexpression of the interferon response is correlated with a worse (ECMO/Fatal) outcome and an increased viral load and proinflammatory cytokines in the early-acute-phase. This report provides insights into the differences in innate immune activation between severe patients that associates with different clinical outcomes, using a non-biased approximation.

**Author Summary:** Hantavirus are rodent-borne zoonotic pathogens that when transmitted to humans cause two diseases: hantavirus renal syndrome in Europe and Asia, and hantavirus cardiopulmonary syndrome (HCPS) in the Americas. The latter, the goal of this work, is a highly lethal disease with a case fatality rate of 30%. Moreover no specific treatment or vaccine is available for this disease. In this study, we analyzed hospitalized HCPS patients with severe disease, to understand how they respond to hantavirus infection. We used a method that can measure every mRNA that is being transcribed in one moment (transcriptome analysis) and thus provide an accurate idea of how cells (specifically peripheral blood mononuclear cells) are responding to infection. The knowledge gained in this study helps us further understand the pathogenesis of this disease and might help us to design specific therapies to treat it.

## Introduction

*Orthohantavirus* (known as hantaviruses) is a family of single-stranded, enveloped, negative-sense RNA viruses that belong to the *Hantaviridae* family of the order *Bunyavirales[1]*. Pathogenic Orthohantaviruses can cause two severe human zoonotic diseases: hemorrhagic fever with renal syndrome (HFRS) in Europe and Asia and hantavirus cardiopulmonary syndrome (HCPS) on the American continent, with a case fatality rate up to 30% [2]. *Andes orthohantavirus* (ANDV) is one of South American HCPS causing hantavirus and the only hantavirus that is known to transmit from person to person [3,4]. After an individual gets infected, the incubation period lasts from seven to 42 days [5,6]. Then a prodromal phase of three to six days starts characterized by nonspecific symptoms including fever, headache and myalgia. Patients may progress to the cardiopulmonary phase which in severe disease is characterized by respiratory symptoms that start with dry cough and evolve to respiratory failure needing mechanical ventilation due to a massive capillary leak into the lung interstitium [7,8]. Severe patients present with circulatory shock and cardiovascular depression that require use of vasoactive drugs [7,9]. Since there is not a vaccine for this deadly disease nor a specific treatment, treatment is based on critical care support. A proportion of these severe patients have a more critical illness that needs extracorporeal membrane oxygenation (ECMO) support and about one third die.

A common hallmark of HCPS is the increased immune activation and vascular permeability that develops during the cardiopulmonary phase producing pulmonary edema and cardiogenic shock in the most severe cases [7]. Viremia is present during the incubation period for up to two weeks before onset of symptoms or appearance of IgM and IgG antibodies [3]. Orthohantaviruses systemically infect and replicate predominantly in endothelial cells (ECs) [10,11], but can also infect epithelial and immune cells such as monocytes/macrophages and dendritic cells [10,12–18]. Hantaviruses do not cause a cytopathic effect [10,11] suggesting that their pathogenic effects are caused rather by the host immune response during infection, which includes a cytokine storm [19]. Increased serum levels of proinflammatory cytokines, such as IL-6, IFN-γ and TNF-α, along with high infiltration of mononuclear cells, mainly CD4^+^ and CD8^+^ T lymphocytes in the lung, are associated with a severe outcome [20–22].

The innate immune response, which is the first reaction to a viral infection, could be one crucial step that modifies how the infected individual will respond to the virus. The regulation of the complement system, a significant component of innate immunity, could also influence the clinical course since deletion in regulatory genes (CFHR1 and CFHR3) have been found to be more frequent in severe HCPS patients [23].

The role of the host’s immune responses in the progression of HCPS and their association with severity remains elusive. In this study, we applied a non-biased, transcriptome approach combined with clinical laboratory data in a cohort of ANDV-infected patients that developed a severe clinical course to explore the factors associated with HCPS severity.

This report provides a comprehensive description of innate immune programs active in PBMCs during acute HCPS patients. Increased type I IFN response, specifically the overexpression of interferon-induced genes (ISGs), is associated with a more severe outcome, defined as patients requiring ECMO or died, and to increased viral load in Peripheral Blood Mononuclear Cells (PBMCs) and increased proinflammatory cytokines levels in serum.

## Results

### Characterization of studied participants

Twelve patients, nine males and three females, with severe HCPS were analyzed in this study. They had a median age of 27, nine being of European ancestry and three with Amerindian ancestry. The 12 HCPS patients were further reclassified into two groups according to their severity: MV+VD (*n*=5) were those who received mechanical ventilation (MV) and vasoactive drugs (VD) as treatment, and ECMO/Fatal (n=7) were those patients that also received ECMO as a treatment or died. The healthy control group (n=9) consisted of four females and five males with a median age of 25 years. There was no difference between groups when comparing demographic data (Table 1). Clinical data including days of oxygen requirement, vasoactive drugs, mechanical ventilation, ECMO treatment, and hospitalization days were available for the cohort (Table 1). These values were similar between study groups except for the days in mechanical ventilation. White blood cells and platelet count, ANDV viral load in PBMCs, together with sub-population immunophenotyping of the earliest measurement (up to 48 hours after onset of cardiopulmonary symptoms) were also compared for these groups. There was no difference between groups on viral load, total leukocyte number, or neutrophil percentage, although a tendency of increase in viral load and a decrease in leukocyte count was observed in the ECMO/Fatal patients (Table 1). A significant difference was found in platelet count in the two HCPS groups compared to healthy controls, as previously described [24]. The blood immune cell populations were evaluated by flow cytometry in the first sample from each patient as described in the methods section. Although no significant difference was found, a trend towards an increase in T helper cells, CD56^Bright^ NK cells and NKT cells, and a tendency for a decrease in cytotoxic T cells, NK cells, and B cells was observed in ECMO/Fatal patients compared to MV+VD ones.

**Table 1.** Demographics, clinical and blood cells immunophenotypes of HCPS patients and healthy controls.

|  | HCPS – Severe Patients |  | Healthy Controls | P value |
| --- | --- | --- | --- | --- |
|  | MV+VD | ECMO/Fatal |  |  |
| No. of patients | 5 | 7 | 9 |  |
| Age (years),<br>Median (25 to 75%<br>IQR) | 22 (10.0-26.50) | 34 (15-36) | 25 (17.0-40.50) | 0.501<sup>a</sup> |
| Gender, n (%) |  |  |  |  |
| Male | 5 (100) | 4 (44.4) | 5 (55.6) | 0.193<sup>b</sup> |
| Female | 0 | 3 (37.5) | 4 (44.4) |  |
| Ethnicity, n (%) |  |  |  |  |
| European | 4 (80) | 5 (71.4) | 8 (88.9) | 0.676<sup>b</sup> |
| Amerindian | 1 (20) | 2 (28.6) | 1 (11.1) |  |
| Days of prodromal<br>symptoms,<br>Median (25 to 75%<br>IQR) | 6 (4.5-7.0) | 6 (5.0-7.0) | NA | 1.000<sup>c</sup> |
| Days of Oxygen<br>requirement | 6 (2.0-7.5) | 10 (4.0-19.0) | NA | 0.149<sup>c</sup> |
| Days of VD duration | 3 (2.0-3.5) | 7 (1.0-9.0) | NA | 0.202<sup>c</sup> |
| Days of MV duration | 4 (3.0-5.5) | 9 (4.0-16.0) | NA | 0.106<sup>c</sup> |
| Days of ECMO | 0 | 4 (0-6.0) | NA | 0.048<sup>c</sup> |
| Days of Hospitalization | 13 (9.5-21.5) | 19 (4.0-59.0) | NA | 0.876<sup>c</sup> |
| **Laboratory exams<sup>d</sup>** |  |  |  |  |
| ANDV Viral Load<br>(copies/ng RNA) | 319.50<br>(82-557) | 616<br>(285.5-3368.25) | NA | 0.286<sup>c</sup> |
| Platelets (cells/mm<sup>3</sup>) | 44500<br>(33250-51250) | 79000<br>(47750-154500) | 236000<br>(201500-253000) | 0.002<sup>a</sup> |
| Leukocytes (cells/mm<sup>3</sup>) | 14900<br>(10000-22300) | 10850<br>(7175-15425) | 6400<br>(5725-7575) | 0.085<sup>a</sup> |
| Neutrophils % | 51 (47-60.47) | 63.8 (37.03-81) | 53 (51-61) | 0.732<sup>a</sup> |
| **Blood cell immunophenotype\*** |  |  |  |  |
| **T cell panel**<br>**(cells/mm<sup>3</sup>)** |  |  |  |  |
| CD3+ cells | 734.97<br>(568.2-2384.37) | 1216.39<br>(443.99-3390.33) | 1234.33<br>(1018.27-1575.23) | 0.866<sup>a</sup> |
| CD4+ T helper cells | 130.08<br>(114.87-419.53) | 481.82<br>(197.12-1920.69) | 926.42<br>(692.69-1308.09) | 0.90<sup>a</sup> |
| CD8+ cytotoxic T cells | 633.95<br>(452.5-1977.55) | 736.34<br>(246.51-1194.27) | 302.99<br>(262.94-336.84) | 0.297<sup>a</sup> |
| **NK cell panel**<br>**(cells/mm<sup>3</sup>)** |  |  |  |  |
| CD56+ NK cells | 83.85<br>(60.01-199.09) | 462.45<br>(110.19-1243.82) | 290.96<br>(191.72-763.20) | 0.258<sup>a</sup> |
| CD3- CD56+<br>NK Bright cells | 2.26<br>(1.62-8.40) | 45.97<br>(7.73-132.95) | 17.52<br>(10.11-24.23) | 0.098 <sup>a</sup> |
| CD3+ CD56+<br>NKT cells | 67.39<br>(37.92-82.22) | 96.73<br>(19.76-117.53) | 42.62<br>(30.10-141.45) | 0.753 <sup>c</sup> |
| **B cell panel**<br>**(cells/mm<sup>3</sup>)** |  |  |  |  |
| CD19+ LB cells | 122.85<br>(94.18-411.87) | 169<br>(84.29-731.69) | 364.27<br>(211.37-421.74) | 0.508 <sup>c</sup> |

Abbreviations: HCPS, hantavirus cardiopulmonary syndrome; NA, not applicable; VD, vasoactive drugs; MV, mechanical ventilation; ECMO, extracorporeal membrane oxygenation; LOS, length of stay; n, number; LB, B lymphocytes. \*Data shown represent the median (25 to 75% IQR) of the first sample from 12 HCPS, 5 VM+VD, 7 ECMO/Fatal and 9 Healthy controls; <sup>a</sup> Kruskal-Wallis test and <sup>b</sup> Chi-square test were used for comparisons between VM+VD, ECMO/Fatal, and healthy controls; <sup>c</sup> Mann-Whitney test was used for comparisons between VM+VD and ECMO/Fatal. <sup>d</sup> Data shown represents the median (25 to 75% IQR) of the first sample of each patient (Day 0-2/Early acute).

The complete clinical course of the 12 HCPS patients in this study along with samples obtained is represented in Fig 1. The prodromic phase with non-specific symptoms varied between three to ten days, and day 0 represents the onset of cardiopulmonary symptoms. Blood samples were collected during the acute phase (early acute between the days 0-2 and late acute samples between days 5-6 after the onset of cardiopulmonary symptoms). One last blood sample was collected in the convalescent-phase (day >60 after cardiopulmonary symptoms onset). Patients that only required mechanical ventilation and vasoactive drugs during their hospitalization were grouped in the MV+VD patients group. Patients requiring ECMO or that died were grouped together in the more severe ECMO/Fatal group of patients.

**Fig. 1.**
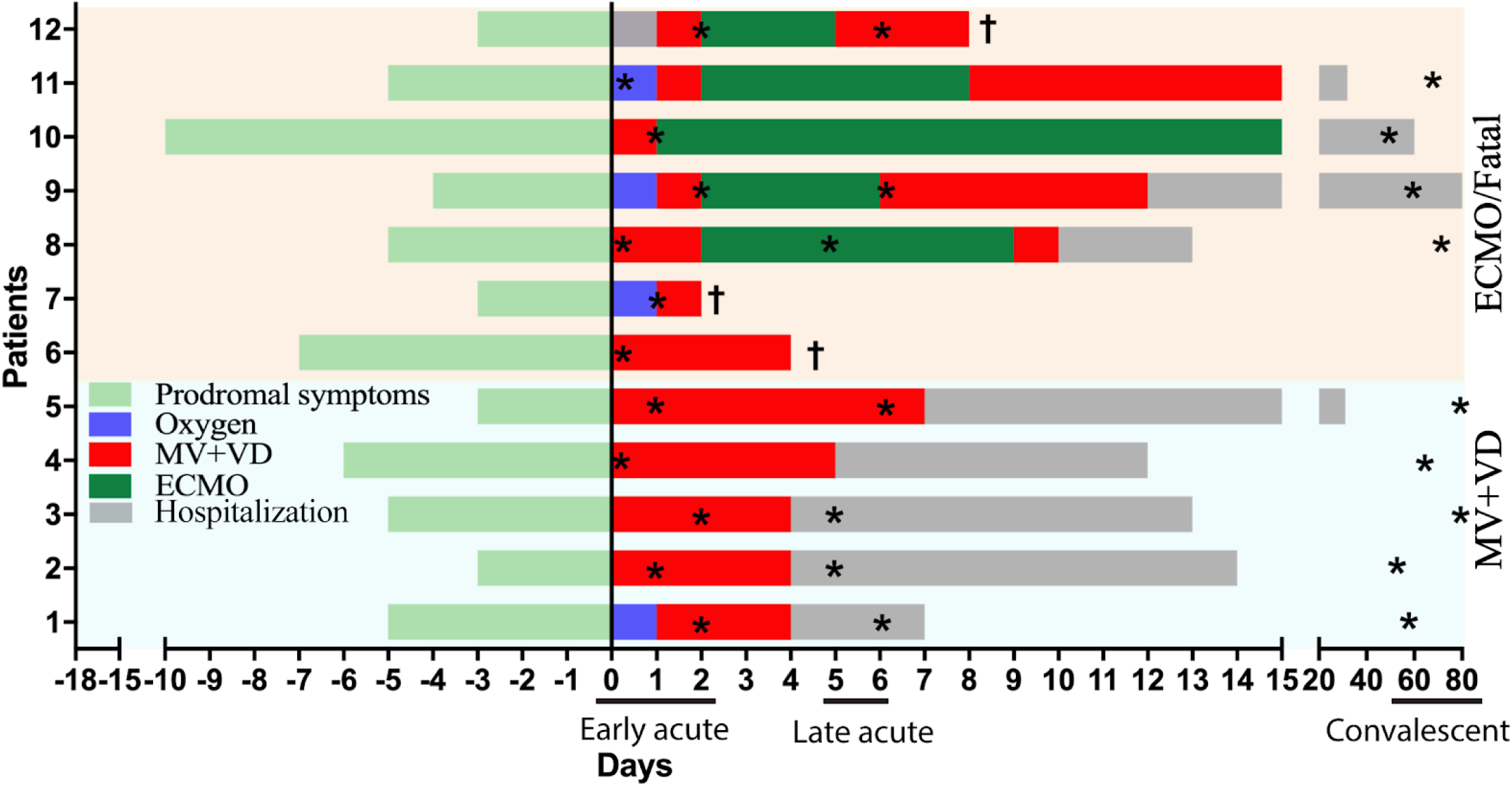
Clinical course of severe HCPS patients. Y axis is patient ID, X axis are the days of evolution, with a highlight on disease timeline: early-acute, late-acute or convalescent phase. Light green bars are the days of onset of nonspecific symptoms. Day 0 represents onset of cardiopulmonary symptoms. Blue bars represent the days when patients required supplementary oxygen. Red bars represent days when patients developed HCPS, requiring vasoactive drugs and mechanical ventilation as treatment. Dark green bars represent patients requiring ECMO. Patients that required ECMO or died were sub-cathegorize to the ECMO/Fatal group. Less severe patients that required vasoactive drugs and mechanical ventilation were sub-cathegorize to VM+VD patients. Grey bars are the hospitalization days to recover. *samples taken, † patients who died. VD, vasoactive drugs; MV, mechanical ventilation; ECMO, extracorporeal membrane oxygenation.

### PBMC transcriptome profile of HCPS patients by severity and clinical phase

Total PBMC samples sequenced and analyzed are detailed in S1 Table. Briefly, after quality control filters, 16,187 genes were included in subsequent analyses. First, a multidimensional scaling (MDS) was performed to evaluate the global gene expression patterns between the samples to determine which variable explained the difference in the data. In MDS analysis two groups were observed: one integrated by healthy controls and convalescent-phase subjects and another group of acute-phase patients (Fig 2A). These results show that the greatest variability in subject’s gene expression was produced by the infection itself. Interestingly, there was no clear separation between MV+VD and ECMO/Fatal patients in early or late-acute-phase.

**Fig. 2.**
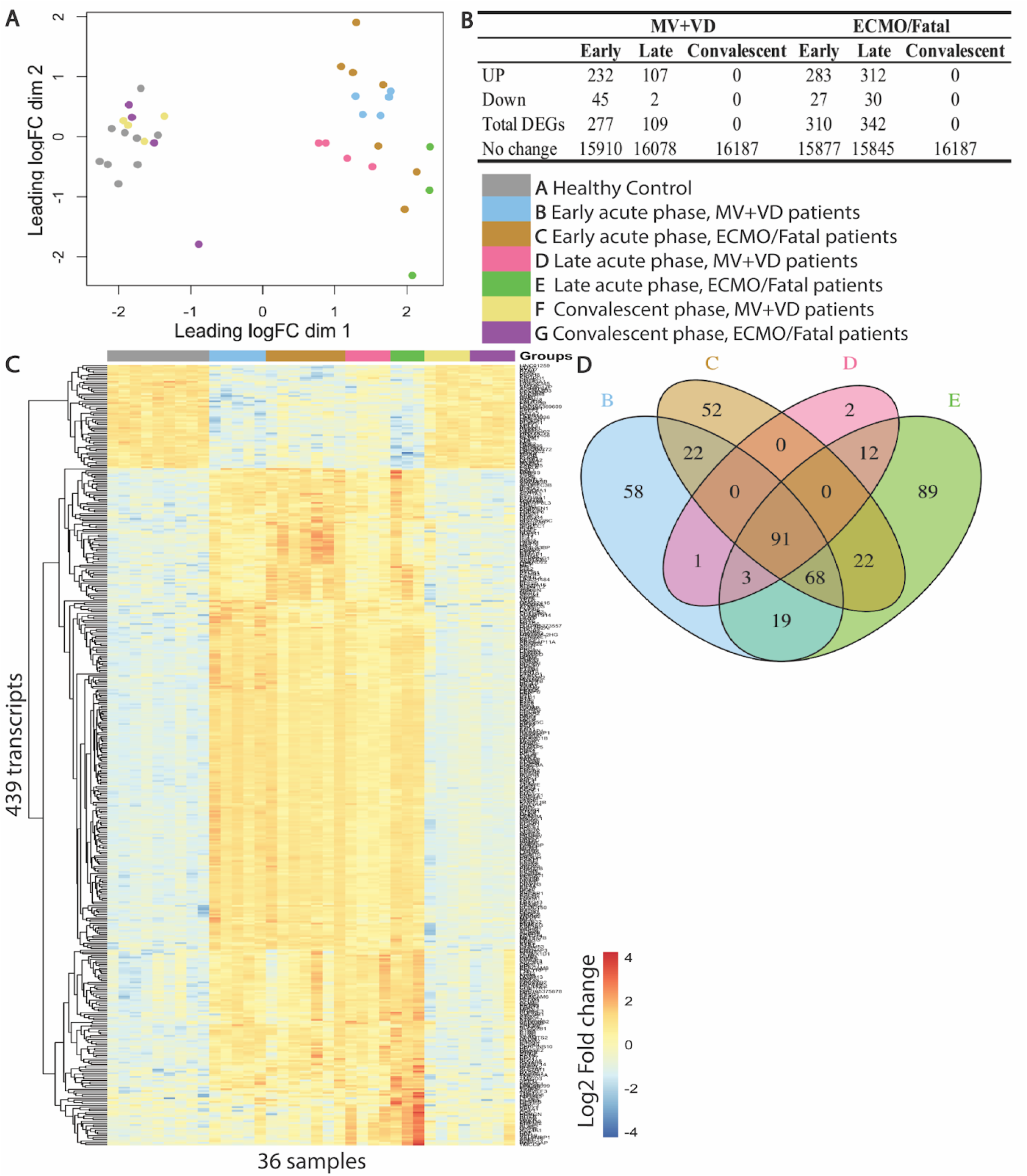
Transcriptional PBMC signature of HCPS patients by severity. **(A)** HCPS patients and healthy controls were divided in seven study groups from A to G (study groups inside figure legend). Transcriptional profile was graphed as a multidimensional scaling (MDS) plot, where X and Y axis represents the variability in global expression profile of the samples and each dot is a patient with the corresponding colour of the study group. **(B)** Number of differentially expressed genes (DEGs) between MV+VD and ECMO/Fatal HCPS patients in an early-late-convalescent phase versus healthy controls with adjusted p-value of 0.05 (FDR of 5 %) and exhibiting at least 2 log2 fold change (log2FC > 2) difference in expression levels. DEGs were obtained by comparing each study group of HCPS patients with the healthy control group. **(C)** Transcriptional signatures of DEGs of healthy controls, MV+VD and ECMO/Fatal HCPS patients in an early and late-acute and convalescent phase. Data are displayed in a heatmap format, where each column represents a patient and each row represents the expression value (log2FC median centered) of each gene between all samples. Red indicates overexpression, blue indicates underexpression, and yellow indicates no difference in expression between samples. The total of DEGs between groups were plotted in the heatmap (439 transcripts). **(D)** The Venn diagram shows the number of shared and unique DEGs between MV+VD and ECMO/Fatal patients in an early and late-acute-phase versus healthy controls.

Differential gene expression was conducted with the limma-voom analysis workflow using Benjamini and Hochberg (BH) correction for multiple testing. Differential expression analysis was performed by comparing samples from MV+VD and ECMO/Fatal HCPS patients in an early-acute, late-acute, and convalescent-phase with healthy controls. The total of differentially expressed genes (DEGs) in the MV+VD group was 277 in early and 109 in late-acute-phase; and in ECMO/Fatal group DEGs were 310 in early and 342 in late-acute-phase (Fig 2B). IFI27 was the most overexpressed gene with highest Log2FC and lowest p-value in both groups of severe patients during the acute phase. Interestingly, proinflammatory cytokines, such as IL-6 and TNF-ɑ, that are known to be increased in serum of severe HCPS patients, were not differentially expressed in MV+VD or ECMO/Fatal patients at any time when compared to healthy controls. Taken together, these results suggest that the increased serum levels of these cytokines could be a contribution of other cell types, such as endothelial cells and macrophages, rather than PBMCs. There were no statistically significant DEGs in the convalescent-phase when compared with healthy controls (Fig 2B). This suggests that at the convalescent-phase, patients have resolved the disease in terms of level of gene expression. At the level of clinical symptoms, some patients continued to be hospitalized because of ECMO complications, others experienced fatigue and mild respiratory distress at day 60, but the majority had returned to normal life. Fig 2C shows the expression profile of MV+VD and ECMO/Fatal HCPS patients at every time point. The total of 439 DEGs were plotted in the heatmap. ECMO/Fatal patients showed a different signature compared to MV+VD patients during the acute phase in both early and late timepoints. As expected, HCPS patients in the convalescent-phase presented a pattern of expression similar to healthy controls. The number of shared and unique DEGs between MV+VD and ECMO/Fatal patients in an early and late acute-phase are shown in a Venn diagram (Fig 2D). There were 58 genes differentially expressed only in MV+VD patients in an early-acute phase and 2 in a late-acute phase and 52 in ECMO/Fatal patients in an early-acute and 89 in a late-acute phase (Fig 2D). Ninety-one genes were shared throughout all conditions (MV+VD and ECMO/Fatal in an early and late-acute phase) compared to healthy controls (Fig 2D).

### Gene set enrichment analysis by severity and clinical phase

To further define the biological function and pathways associated with a severe clinical course, we applied a gene set enrichment analysis called blood transcriptome module (BTM) as described [25,26]. BTMs are groups of genes that share a similar function, where BTM up- and downregulation is defined by the percentage of genes within each module that are differentially expressed. Activation of modules was tested comparing each study group to healthy controls and using the FDR-ranked lists of DEGs generated by limma and applying the tmod test. The BTMs with a *p-value* ≤ 0.001 were considered significant. When HCPS patients were analyzed together, a strong upregulation of Modules that group genes with functions in cell cycle, DNA repair, mitosis was observed (data not shown), which can be further observed when patients are re-classified in MV+VD and ECMO/Fatal groups (Fig 3). Moreover, this response is still active during the late-acute-phase but returns to normal values during convalescence. The enrichment observed in these pathways points to the immune activation that is taking place in both groups of severe patients and the lymphoid proliferation that is taking place during the acute-phase of the disease. On the other hand, when all HCPS patients were analyzed together, modules that grouped genes from the innate immune response were enriched during the early phase of the acute disease (data not shown). When these severe patients were reclassified as MV+VD and ECMO/Fatal, this association was only seen in the more severe group (ECMO/Fatal) during the early-acute phase (Fig 3, marked with arrows). These BTMs include a greater activation in antiviral IFN signature, IFN I signalling and innate antiviral response, activation in B cells and plasma cells and BTMS of activated dendritic cells. The comparison of expression of the significant genes belonging to these innate immune response marked modules, are shown in S2 Table. This data suggests that in ANDV-infected patients that develop HCPS, there is a strong innate immune response that is greater in the most severe ECMO/Fatal HCPS group of patients. Interestingly, these more severe ECMO/Fatal patients have an over-representation of innate immune pathways, especially genes participating in the interferon signaling pathway.

**Fig. 3.**
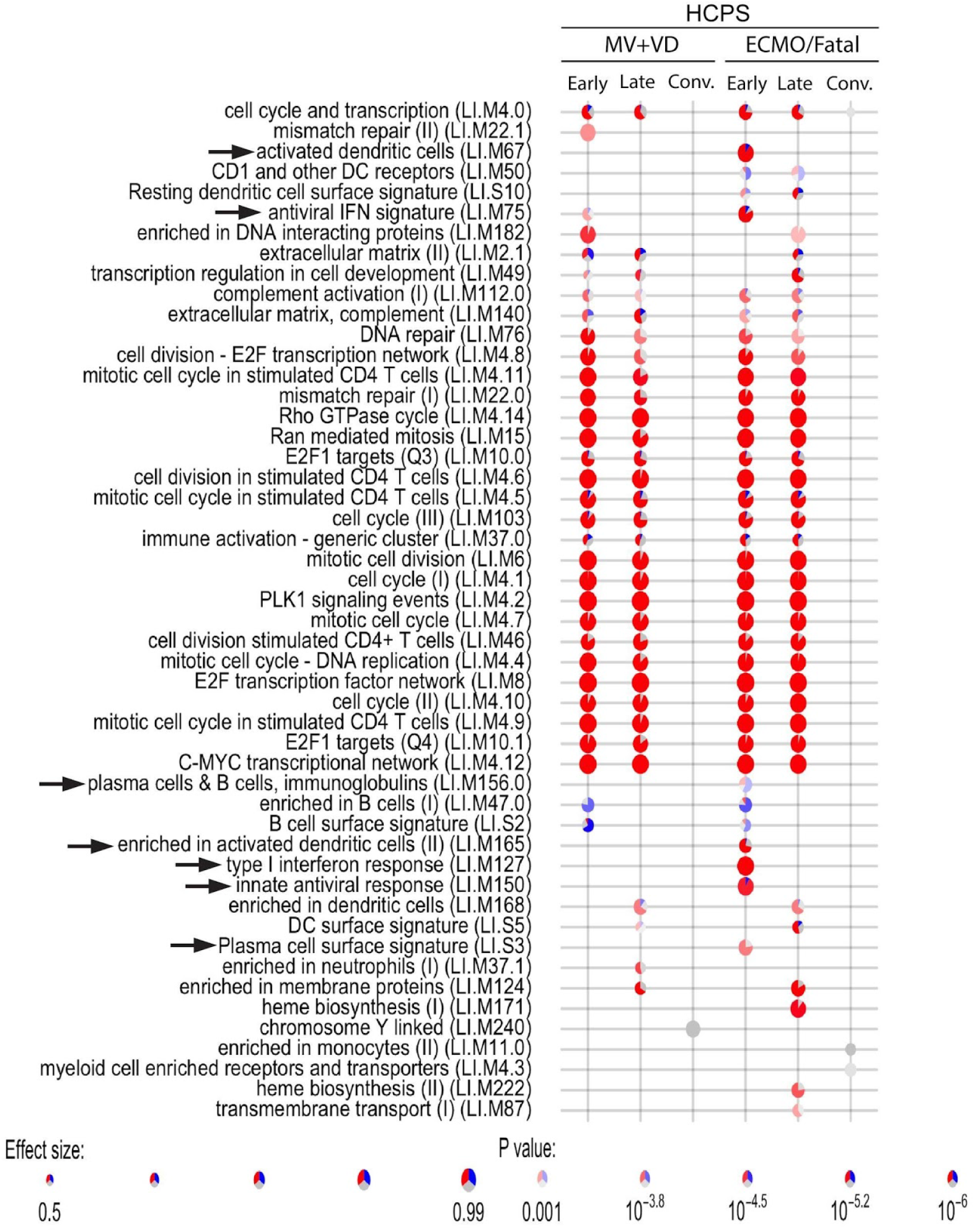
Dynamics of VM+VD and ECMO/Fatal HCPS patient’s response in PBMCs. BTM dot plot of differentially enriched BTMs in VM+VD and ECMO/Fatal HCPS patients, in early-late-convalescent-phases were compared with healthy controls. Y axis has the name and code of each BTM module, and X axis each study group. Activation of modules was tested using the FDR-ranked lists of genes generated by limma and applying the tmod test. Each comparison is in the intersection of lines of the X and Y axis. When a module is statistically significant (p-value ≤ 0.001) it is represented by a pie chart in the intersection in which the proportion of significantly upregulated genes are shown in red and downregulated in blue. The grey portion of the pie represents genes that were not significantly differentially regulated. The effect size (enrichment) is proportional to its size which means that the larger the size of the pie, the greater the module enrichment. The significance of module activation (*p-value*) is proportional to the intensity of the color pie, which means that the greater the color intensity (stronger the red or blue color), the lower the *p-value* and the greater the significance. Modules that were not statistically significant, were not plotted and are seen as the line cross of X and Y axis.

### DEGs in MV+VD and ECMO/Fatal HCPS patients in early-acute phase

Once the functions of all DEGs between MV+VD and ECMO/Fatal patients in an early-late acute phase were described, unique genes for each severity group were analyzed. One of the interests of the study was to understand the factors associated with critical clinical courses at the onset of the cardiopulmonary phase, so further analyses were focused on unique DEGs of the early-acute phase. Transcriptional profiles of 110 genes that include 52 genes that were detected to be uniquely differentially expressed in ECMO/Fatal patients and 58 genes detected in MV+VD patients were analyzed. Fig 4 shows a clear difference in the expression profile of these genes between patients of different severities and healthy controls.

**Fig. 4.**
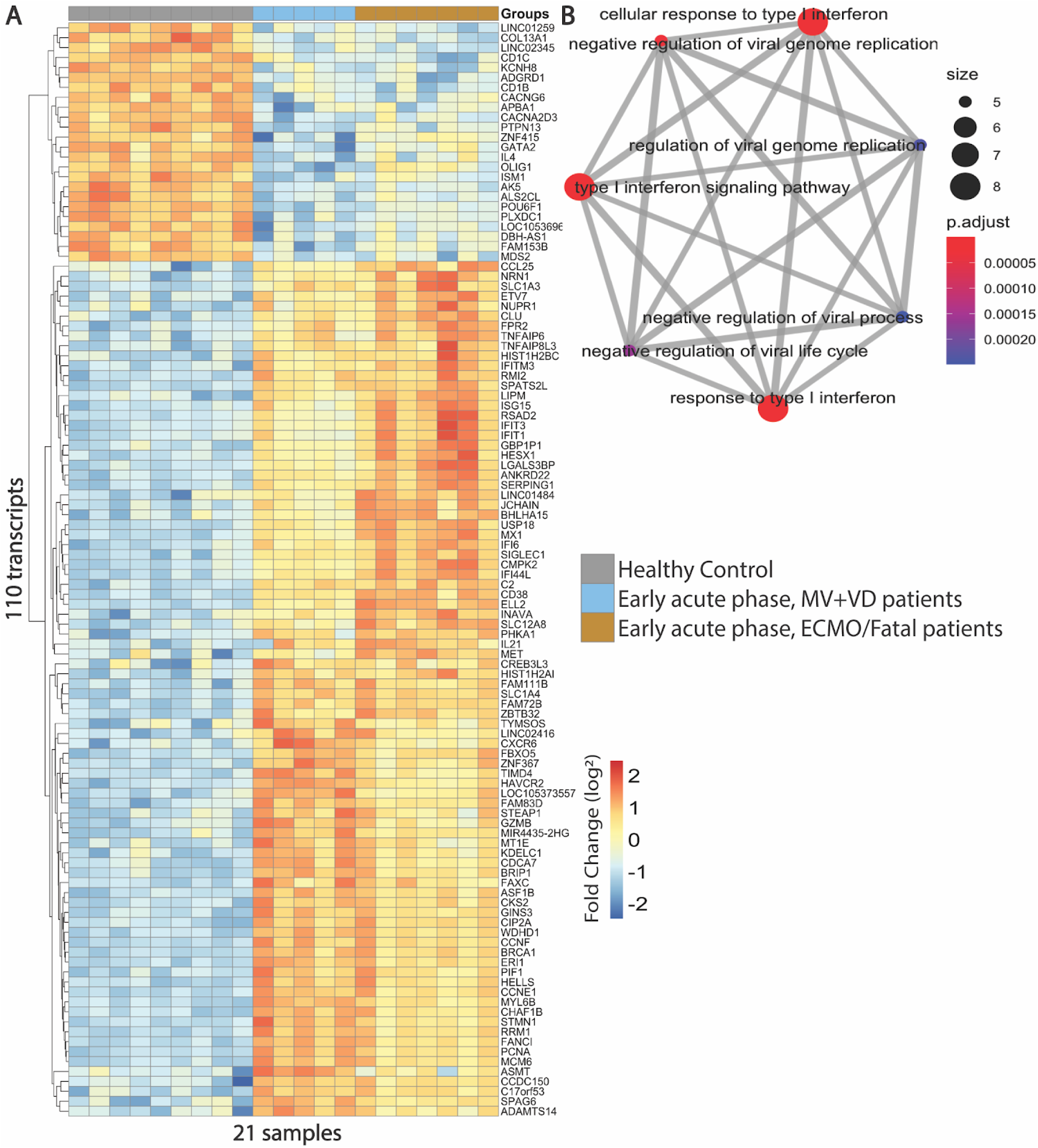
Transcriptional profile of DEGs in VM+VD and ECMO/Fatal patients in an early-acute phase. **(A)**. A total of 110 transcripts are displayed in a heatmap format, where each column represents the gene expression of a patient (five VM+VD, seven ECMO/Fatal and nine healthy controls); Each row represents expression value (log2FC median centered) for each gene between all samples First row corresponds to the colour of the study group (legend inside the figure). Red indicates overexpression, blue indicates underexpression, and yellow indicates no difference in expression between samples. **(B)** Results of biological processes enriched in ECMO/Fatal patients. Gene set enrichment analysis was performed for the 52 exclusive DEG in ECMO/Fatal patients in an early-acute-phase. Node size correlates with gene set size and edge widths indicate the number of shared genes. Red and blue color indicate more and less enrichment significance respectively.

To evaluate the functions of these unique DEGs, a gene set enrichment analysis was performed separately for each group of genes (58 for MV+VD and 52 ECMO/Fatal). This method uses statistical approaches to identify significantly enriched or depleted groups of genes that participate in the same biological process. Thus, the functions which are exclusively altered in each group can be identified.No biological processes were found to be enriched in MV+VD patients compared to healthy controls. However, seven biological processes were enriched in ECMO/Fatal patients. These biological processes were mainly from innate immune response, primarily type I interferon response (Fig 4B). The detail of the genes belonging to these biological processes is in the S3 Table. These results showed that genes associated with innate immune response, especially type I interferon response, were enriched only in ECMO/Fatal patients. This is in concordance to what was found on BTM analysis.

### Correlation between expression of BTM enriched exclusively in ECMO/Fatal patients and clinical laboratory data

To assess whether the expression of the genes from enriched BTMs in ECMO/Fatal patients in an early-acute-phase could impact the severity and clinical laboratory parameters, the expression of these BTMs was correlated with these patients’ laboratory data (Fig 5 and detail in S4 Table). The BTMs associated with interferon response, including antiviral interferon signature (LI.M75), type I interferon response (LI.M127) and innate antiviral response (LI.M150); dendritic cells: activated dendritic cells (LI.M67) and enriched in activated dendritic cells (LI.M165); and plasma cells surface signature (LI.S3), showed a statistically positive correlation with ECMO/Fatal clinical course (Fig 5). The BTMs associated with interferon response (LI.M75, LI.M127, and LI.M150) are positively correlated with viral load, amount of NK cells and TNF-ɑ serum levels. LI.M127 and LI.M150 are also positively correlated with the amount of NKT cells and IL-6 serum levels and negatively correlated with neutrophils percentage. The BTM activated dendritic cells (LI.M67) is positively correlated with NK cells, viral load, amount of NK, and Th cells. Platelets, leukocytes, CD3+, CD8+, CD56 ^Bright^ NK cells did not show any statistically significant correlation with the enriched BTMs. Thus, the overexpression of interferon response genes is correlated with a ECMO/Fatal outcome and an increased viral load in PBMCs and proinflammatory cytokines (IL-6 and TNF-ɑ) serum levels and could be contributing to immunopathogenesis instead of restricting viral replication. Having an increased type I interferon response appears to correlate to poor prognosis in HCPS patients.

**Fig. 5.**
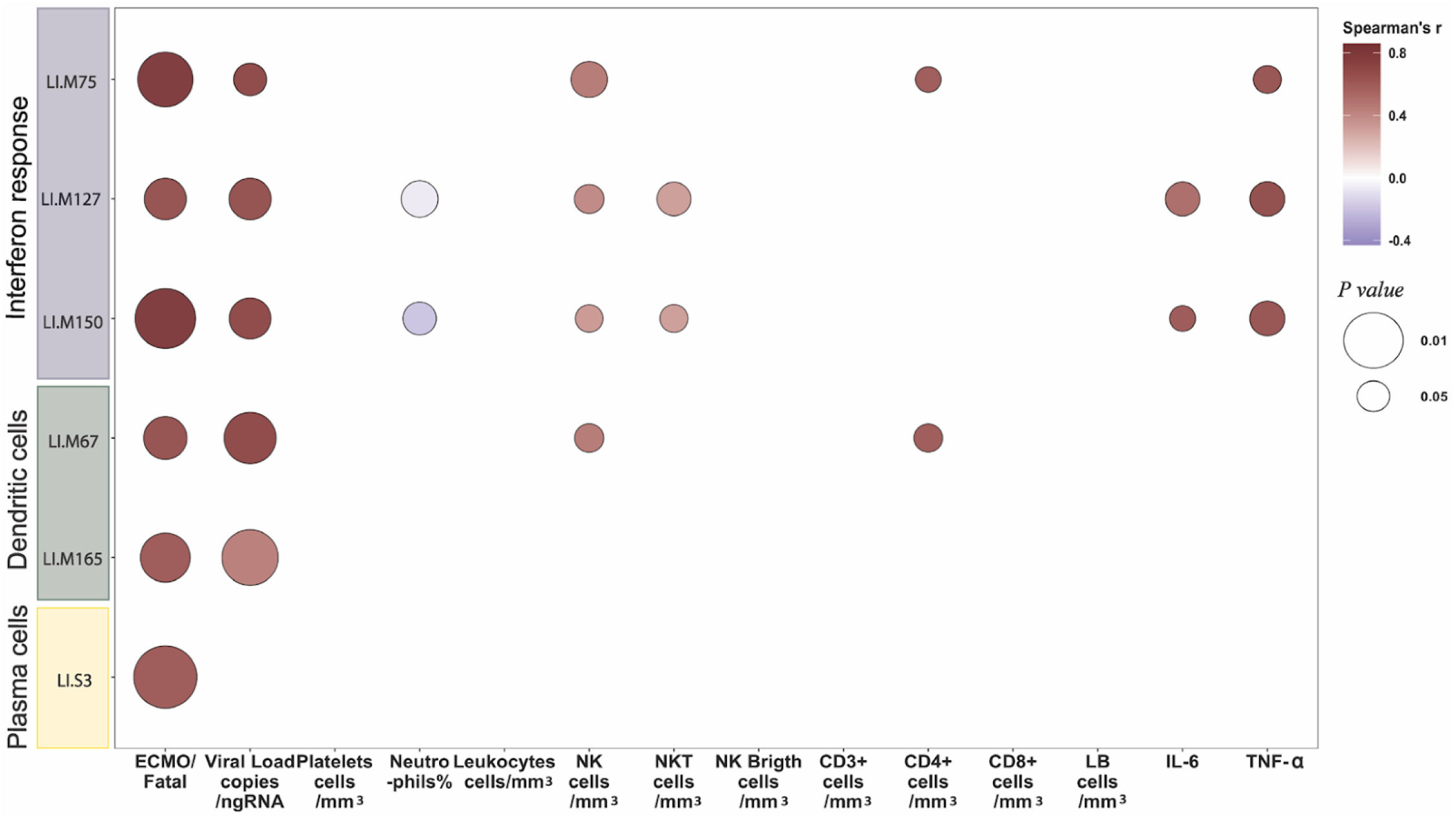
Correlations between transcriptional profile enriched in ECMO/Fatal patients in an early acute response and clinical laboratory data. X axis has clinical, laboratory data and immune profile information Y axis has the code of analyzed BTMs and to which biological process it belongs. Spearman correlation between expression median of genes over- or underexpressed per BTM data and clinical/lab/immune information was performed for VM+VD and ECMO/Fatal patients. The dot in the intersection of X and Y axis shows the correlation, if no dot is present there’s no correlation. The intensity of the color in the dot indicates the strength of the Spearman’s correlation coefficient (red positive correlation and blue negative correlation), and the dot size indicates the significance of the *P-value*. CD3, CD3+ cells; CD4+, T helper cells; CD8+, cytotoxic T cells; LB, B cells; LI.M75, antiviral interferon signature; LI.M127, type I interferon response; LI.M150, innate antiviral response; LI.M67, activated dendritic cells; LI.M165, enriched in activated dendritic cells; LI.S3, plasma cells surface signature.

## Discussion

The host immune response is an important mechanism in the pathogenesis of HCPS. An exaggerated and uncontrolled activation of the innate and adaptive immune response is associated with severity [20,27–31]. In this study, we applied a transcriptome approach combined with clinical laboratory data to gain a better insight into factors associated with HCPS critical or fatal clinical course.

The human innate immune response, particularly type-I IFN response, is a highly robust and effective first line of defense against virus invasion. In infected and neighboring cells, type I IFNs induce the expression of IFN-stimulated genes (ISGs), the products of which initiate an intracellular antimicrobial program that limits the spread of infectious agents [32,33]. Failure to mount an effective IFN response against the virus leads to systemic infection, while excessive IFN production leads to pathogenicity, severe symptoms, or even fatality [34–36].

Our findings demonstrated that PBMCs transcriptome from the more severe ECMO/Fatal HCPS group of patients have a greater activation of the innate immune response in the earliest stages of the cardiopulmonary phase than in less severe MV+VD patients. BTMs associated with IFN response, dendritic, and plasma cells were enriched in the ECMO/Fatal patients between 0 to 2 days after cardiopulmonary symptoms started. They also had upregulated genes associated with type I interferon response, whose main function is to negatively regulate viral replication (i.e., USP18, ISG15, IFIT3, IFIT1, MX1, IF6, IFI44L, IFITM3 and Siglec-1). In other viral infections, such as Coronavirus disease 19 (COVID-19), severe patients have recently been characterized of having an up-regulation of various ISGs, including ISG15, IFITM1/2/3, and ISG20[31]; so, these unregulated type I IFN responses could be an important hallmark for severity in viral respiratory diseases. Interestingly, the BTMs enriched in the ECMO/fatal HCPS patients associated with type I IFN response (LI.M75, LI.M127, and LI.M150) were positively correlated with high viral load, proinflammatory cytokines (IL-6 and TNF-ɑ) serum levels and ECMO/Fatal clinical course. BTM associated with dendritic cells (LI.M67, LI.M165) were also positively correlated with viral load and ECMO/Fatal clinical course. This means that the higher the expression of type I IFN genes, the greater the viral load and proinflammatory response and consequently greater illness severity. Because the overexpression of interferon response genes is associated with a critical and fatal outcome, an increase in viral load in PBMCs and an increase in proinflammatory cytokines serum levels, it could rather be contributing to immunopathogenesis rather than restricting viral replication in these patients.

It has been known for decades that viruses evolve different strategies to escape the type I IFN response in order to replicate and disseminate successfully in their hosts [37]. *In vitro* experiments have shown that pathogenic hantaviruses can block early IFN responses, which favor an early viral replication in target cells, but later induce high-level ISG responses (1–4 days after infection) [13,38,39]. These observations correlate with HCPS natural history in which there is a long incubation period that can last up to 49 days, and viremia can be detected during this period up to 15 days before symptoms onset, suggesting a long escape of the virus to IFN response.

Moreover, *in vivo* experiments have shown that the treatment with IFN-β could increase the survival of Hantaan (HTNV) infected mice only if performed less than 24 hours after infection [40]. In this study we have shown that despite a strong interferon response in the ECMO/fatal HCPS patients, there is no correlation with a decrease in viral load. The findings of this study suggest that insensitivity to the interferon response occurs in ECMO/fatal HCPS patients and correlates with poor prognosis in HCPS patients. The molecular mechanisms by which hantaviruses become insensitive to the interferon response, specifically how they evade the response to ISGs once the infection is already established, is not yet known and should be studied further.

The mechanisms by which type I interferon response promotes severity in viral infections remains controversial and elusive. A common theme is emerging in which type I IFN has the potential to over activate the immune system during acute viral infection. Besides the induction of ISGs, type I IFN also induces secretion of cytokines and chemokines and activation of pathways that allow the clearance of infected cells [41,42]. Therefore, the induction of proinflammatory cytokines or chemokines or activation of apoptosis-inducing pathways to clear virally infected cells, which were designed to be protective, can lead to tissue damage with serious consequences to the host [41,43]. HCPS is characterized by an exacerbated immune response that includes a “cytokine storm” that contributes to pathogenesis. In this sense increased serum levels of proinflammatory cytokines, such as IL-6, IFN-γ and TNF-ɑ are associated with a severe HCPS outcome [20,22,44]. We found that the overexpression of ISGs is positively correlated with proinflammatory cytokines (IL-6 and TNF-ɑ) serum levels. This suggests that increased type I IFN response could be contributing with the increase of proinflammatory response in the ECMO/fatal HCPS patients. Although these proinflammatory cytokines were not differentially expressed in severe or ECMO/Fatal patients when compared with healthy controls, the increased serum levels of these markers could be a contribution from other cell types, such as endothelial cells or macrophages, rather than PBMCs. Thus, the overregulation of ISGs in PBMCs could be contributing to immunopathology altering inflammatory pathways in a paracrine way.

Our results provide insights into the dynamics of early and late PBMCs immune responses in HCPS patients and factors associated with ECMO/fatal clinical course. We found that the overexpression of type I IFN genes are positively correlated with increased viral load in PBMCs and proinflammatory cytokines and could contribute to immunopathogenesis rather than restricting viral replication in HCPS patients. Then, hantavirus IFN insensitivity together with exacerbated proinflammatory response induced by increased IFN response could contribute to ECMO/fatal HCPS outcome (Fig 6). These results suggest that treatment with interferon could worsen the HCPS clinical course. Future studies should investigate the damaging effects of type I IFN response to better understand the immunopathogenesis of HCPS.

**Fig. 6.**
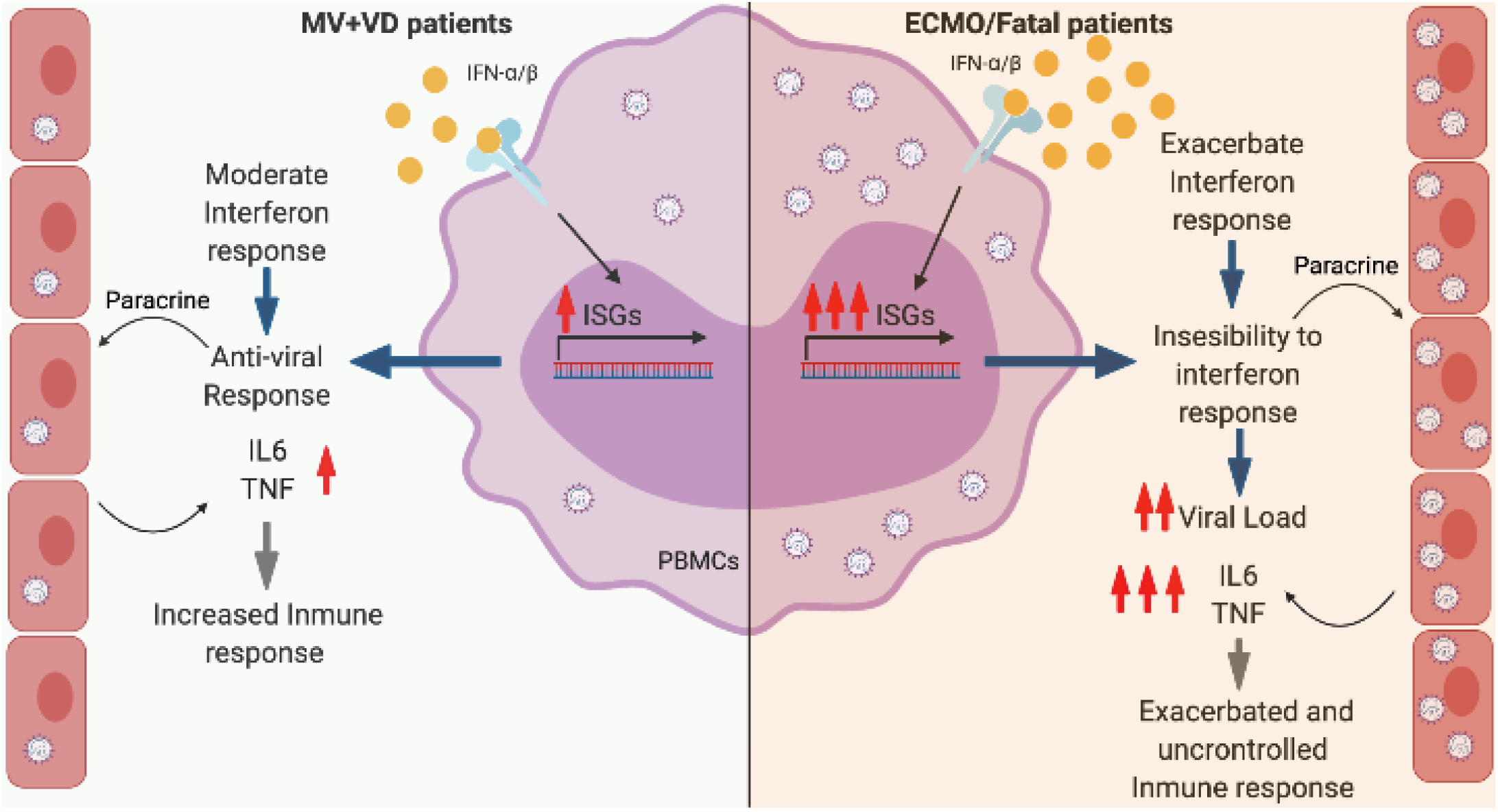
Increased type I IFN is a hallmark of disease severity in HCPS patients. After infection is established in an early-acute-phase, type I IFN response is expressively activated and insensitivity to IFN can happen in the more severe patients that are treated with ECMO or die. Also, other cells such as endothelial cells produce high levels of proinflammatory cytokines that contribute to exacerbated immune response in more severe patients. Less severe patients present a moderate interferon response which leads to an antiviral and controlled immune response. Created with Biorender.

## Materials and methods

### Ethics statement

This study was approved by the ethics committee of each participating institution, Clínica Alemana Universidad del Desarrollo (UDD) IRB4858, FWA8639, January 2016; Hospital Clínico UC, IRB2886, FWA4080, July 2016; Hospital Base Valdivia IRB1914 FWA 3412, January 2017, Hospital Base Osorno IRB1914 FWA 3412, January 2017; Hospital Regional Puerto Montt, IRB1914, FWA 3412, January 2016, Instituto Nacional del Tórax, April 11 2017. Enrolled subjects provided written informed consent, their parents, or legal guardians. Clinical data and stored samples were anonymized to ensure confidentiality.

### Study design

Subjects were recruited in six research centers from central and southern regions of Chile between March, 2017, and June 2018. Hospitalized subjects with a suspected or confirmed diagnosis of ANDV infection by serology or RT-qPCR were invited to participate. Exclusion criteria included patients connected to extracorporeal membrane oxygenation (ECMO) at enrollment. A total of 24 HCPS patients were enrolled, eight were excluded for these analyses: one co-infected with Epstein-Barr virus, another one negatively diagnosed for HCPS, patients with a mild clinical course (not requiring vasoactive drugs or mechanical ventilation) or that did not complete follow up. A group of healthy subjects (controls) were also enrolled (n=9), and exclusion criteria included the presence of chronic disease, pregnancy or any signs and symptoms of an acute infection in the two weeks prior to enrollment. A schematic diagram of enrolled and analyzed patients is available in S1 Fig. The 12 severe HCPS patients analyzed were defined as those who develop cardiopulmonary failure and required vasoactive drugs and mechanical ventilation. Severe HCPS patients were further reclassified as MV+VD (n=5) for those who received only mechanical ventilation and vasoactive drugs as treatment and ECMO/Fatal (n=7) for those who also received ECMO as a treatment or died. The subjects were followed for sixty days post-enrollment. Two blood samples were collected during the acute phase and one sample in convalescent-phase (at least 60 days after onset of cardiopulmonary symptoms). A single sample was taken for healthy controls.

### Sample preparation and RNA-seq

The PBMCs were separated by Ficoll-paque (Sigma Aldrich) following the manufacturer’s specifications. Aliquots of 1-3×10^6^ PBMCs/mL were prepared and the RNA was stabilized with RNAprotect cell reagent (QIAGEN) and frozen at -80°C. The RNA was extracted using the RNAeasy kit (Qiagen) following the manufacturer’s specifications. The quality was assessed using the Agilent 2200 TapeStation. All samples exhibited RNA integrity numbers (RIN) greater than 6. Aliquots of 350 ng were prepared, stored at -80°C and shipped on dry ice to the Broad Institute (Boston, USA) for RNA sequencing. RNA-seq libraries were prepared with Illumina Truseq Stranded mRNA chemistry. Libraries were sequenced in multiplex on the Illumina HiSeq 2500 platform in 50M reads in pairs.

### Viral Load and multiplex

For the viral load determination, total nucleic acids were extracted from the buffy coat obtained from 5mL of peripheral blood using High Pure Viral Nucleic Acid Kit (Roche) following the manufacturer’s specifications. ANDV virus infections were determined by reverse transcription-polymerase chain reaction (RT-qPCR) using primers from the S segment as described previously[45] and the number of viral copies per total ng RNA was calculated. The serum levels of proinflammatory cytokines (IL-6, TNF-ɑ) were measured using a MILLIPLEX map (HCYTOMAG-60K, Merk) following the manufacturer’s instructions.

### Transcriptome and BTM analysis

A total of 37 samples from 12 HCPS patients (five MV+VD and seven ECMO/Fatal in an early, late and convalescent-phase) and nine healthy controls were sequenced (S1 Table). Quality control of the sequenced reads was performed with FastQC. The low-quality sequences (score <30) and Illumina adapters were excluded using trimmomatic (v0.38)[46]. Reads were mapped to the human genome reference (hg37) using the STAR (v2.7.0d)[47] alignment tool. Read counts per gene were quantified against Ensembl (hg37) transcript reference annotations using featureCounts (v1.6.2)[48]. Analysis was conducted within the Jupyter notebook using IRKernel (v0.8.15) statistical framework. First, filters to remove samples with less than 10 million aligned reads were applied. Only one sample of convalescent-phase from Severe type 1 patient did not meet these quality criteria and was excluded from the analyzes. 36 sequenced samples passed filter and were included in the following analysis (S1 Table). Second, RNA-Seq read counts were scaled and normalized by differences in library size between samples using log2-counts per million (log-CPM). Expressed genes were defined as genes with greater than one log-CPM in at least four samples. Genes that did not meet this parameter were filtered. 16,187 genes passed the filter. Third, the RNA-Seq read counts were normalized by composition bias which computes normalization factors for comparing between libraries on a relative scale using the Trimmed Mean of M-values (TMM) method[49] and log2 transformed using voom as previously described[50]. log-CPM and TMM normalizations were performed with edgeR package (v3.24.3)[51]. To assess the similarities and dissimilarities in global expression profile between the samples Multi-dimensional scaling (MDS) was performed by limma package (v3.38.9)[52] and plotted by plotMDS function. Differential expression (DE) was conducted with the limma-voom analysis workflow as previously described[50] using Benjamini and Hochberg (BH) correction for multiple testing. DE was performed to compare samples from MV+VD early, late and convalescent; and ECMO/Fatal early, late, and convalescent versus healthy controls. Differentially expressed genes (DEGs) were selected based on an adjusted p-value of 0.05 (FDR of 5 %) and exhibiting at least 2 log2 fold change (log2FC > 2) a difference in expression levels. To identify the exclusive DEGs on each condition and those shared throughout all conditions a Venn diagram was made using the VennDiagram (v1.6.20) package. The gene set enrichment was performed using the clusterProfiler (v3.14.3). Enriched biological processes with an adjusted p-value of 0.01 were considered statistically significant. The heatmaps were scaled (log2FC median centered) and clusterized by row and graphed by pheatmap package (v. 1.0.12). Blood transcriptome modules (BTM) enrichment analyses were conducted with tmod (v.a40)[53] using the tmodLimmaTest function. BTM gene memberships and annotations were obtained from Li et al[25]. BTMs with an adjusted p-value of 0.01 (FDR of 1 %) were considered statistically significant.

### Blood immune cell populations

Blood samples (5 ml) were obtained in acid citrate dextrose (ACD) tubes (BD Vacutainer ACD Solution B; BD, Franklin Lakes, NJ) only on early-acute-phase and processed within 2 to 4 hours of collection. PBMCs were separated by Ficoll-paque (Sigma Aldrich) and stored at -80°C using CryoStor CS10 medium (STEM CELL technologies) until use. Thawed PBMCs were stained with different antibody panels for characterization of NK, T and B cells subpopulations (S5 Table) using BD Biosciences antibodies. PBMCs were rapidly unfrozen and washed with 10% FBS supplemented PBS, centrifuged at 1200 rpm, and resuspended in PBS-2% FBS. PBMCs were counted and 2.5-5.0 × 10^5^ live PBMCs were used for antibody stainings. Stainings were performed at room temperature for 15 minutes protected from light. PBMCs viability was assessed with the Zombie Aqua Fixable Viability Kit (Biolegend® cat. 423102). After that, unbounded antibodies were washed away two times and stained PBMCs were fixed with 200 uL 2% paraformaldehyde. The acquisition was performed in a FACS Canto II flow cytometer (BD Biosciences) and the data was analyzed with FlowJo v10.0.7.

### Statistical analysis

RNASeqPower program package (v3.0.1)[54] was used for sample size calculation, which was developed to estimate the statistical power necessary to identify differentially expressed genes from RNA-seq experiments. To estimate the sample size, the estimated power of 0.8, the coefficient of variation of 0.4, and an effect size of 1.75 were used. Considering these parameters, the number of patients required for differential expression analysis was 9 for each group. For the analysis of the patient’s demographic and clinical characteristics, IBM SPSS Statistics v.26 was used. Shapiro normality test was applied, and the Mann-Whitney test was used for comparisons between two non-paired numeric variables and Kruskal-Wallis for three or more groups without normal distribution, and Chi-square test for qualitative variables. To assess the correlation between BTM expression and clinical data, the expression median of genes from BTMs for each patient and Log2FC were calculated. Spearman correlation was performed in the R program using cor() function and was graphed with gplots (v3.0.1.1).

## Data Availability

FastQ files from transcriptomic analysis are available at PRJNA660433. Any metadata needed must be obtained through an MTA with the authors.

## Acknowledgments

Thank members of the Hantavirus Study Group in Chile who contributed to patient enrollment and follow-up, sample collection and data management are as follows: Catalina Infante, RN, Carolina Henríquez, RN (Departamento de Enfermedades Infecciosas e Inmunología Pediátricas, Pontificia Universidad Católica de Chile); Flavio Carrión, PhD, Rodrigo Pérez, BKin (Clínica Alemana de Santiago Universidad del Desarrollo), Pamela Silva, RN, Angélica Gavilán, MT, Iván Rodríguez, MT, Marina Opazo, MD (Hospital Dr, Guillermo Grant Benavente Concepción); Carola Osorio, MT, Paulina Miranda, RN, (Hospital Base Valdivia) María Paz Blanco, RN, Camila Bolados, RN, Catherine Bosnich, MT, Paulina Cárcamo, MT, Carolina Nuñez, EU (Hospital Puerto Montt Dr. Eduardo Schütz Schroeder); Felipe Vargas, MT, (Hospital Base San José de Osorno);. Thanks Dr. Anne Bliss and Steven Bradfute, for critical reading of the manuscript, and Dr. David Gorkin for critical review of the bioinformatic pipeline. Funding: This research was funded by Fondecyt 1161447, Fondecyt 1201240, Redes 180195, Concurso Interno VRID UDD2018, Facultad de Medicina, Clínica Alemana Universidad del Desarrollo.

## Supporting Information captions

**S1 Fig. Schematic overview of enrolled and analyzed individuals for this study**.

**S1 Table. Number of sequenced samples of HCPS patients and Healthy controls**.

**S2 Table. Significant DEGs belonging to marked BTMs from text Fig 3**

**S3 Table. Gene enrichment analysis: Biological processes and enriched genes in ECMO/Fatal group of patients in early-acute-response, from Fig 4B**

**S4 Table. Correlations between transcriptional profile enriched in ECMO/Fatal patients in an early acute response and clinical data**.

**S5 Table. Cell surface markers used to define WBC populations by flow cytometry immunophenotyping**.

## Financial Disclosure Statement

This research was funded by Agencia Nacional de Investigación y Desarrollo (ANID) Projects Fondecyt 1161447, Fondecyt 1201240, Redes 180195; Concurso Interno VRID UDD2018, Universidad del Desarrollo.

## Competing interests

The authors declare no conflict of interest.

## Related manuscripts

The authors declare there is not a duplicated or related manuscript under consideration or accepted for publication elsewhere.

## Author contributions

Conceptualization C.V., P.V. and G.E.; methodology C.V., L.L. and GE; patient enrollment M.F., N.LC., C.M-V., L.F, M.L.R, J.G., F.A., J.G.,R.L.,J.L.P; formal analysis, G.E. and L.L.; investigation, R.P., E.D.,C.M-V., P.V., A.C.; resources, P.V. and C.V.; data curation, A.C.; writing original draft preparation, G.E, C.V. and L.L., writing review and editing, N.LC., M.C., G.M. and P.V.; project administration, R.P.; funding acquisition, G.M., P.V. and C.V.

## Notes

### Competing Interest Statement

The authors have declared no competing interest.

### Funding Statement

This research was funded by Agencia Nacional de Investigacion y Desarrollo (ANID) Projects Fondecyt 1161447, Fondecyt 1201240, Redes 180195; Concurso Interno VRID UDD2018, Universidad del Desarrollo.

### Author Declarations

Ethics statement This study was approved by the ethics committee of each participating institution, Clinica Alemana Universidad del Desarrollo (UDD) IRB4858, FWA8639, January 2016; Hospital Clinico UC, IRB2886, FWA4080, July 2016; Hospital Base Valdivia IRB1914 FWA 3412, January 2017, Hospital Base Osorno IRB1914 FWA 3412, January 2017; Hospital Regional Puerto Montt, IRB1914, FWA 3412, January 2016, Instituto Nacional del Torax, April 11 2017. Enrolled subjects provided written informed consent, their parents, or legal guardians. Clinical data and stored samples were anonymized to ensure confidentiality.

