## Supplementary material for "Transcriptomic signature on Hantavirus Cardiopulmonary Syndrome patients, reveals an increased interferon response as a hallmark of critically ill patients": Supple Figs and Tables

### SUPPLEMENTARY INFORMATION:

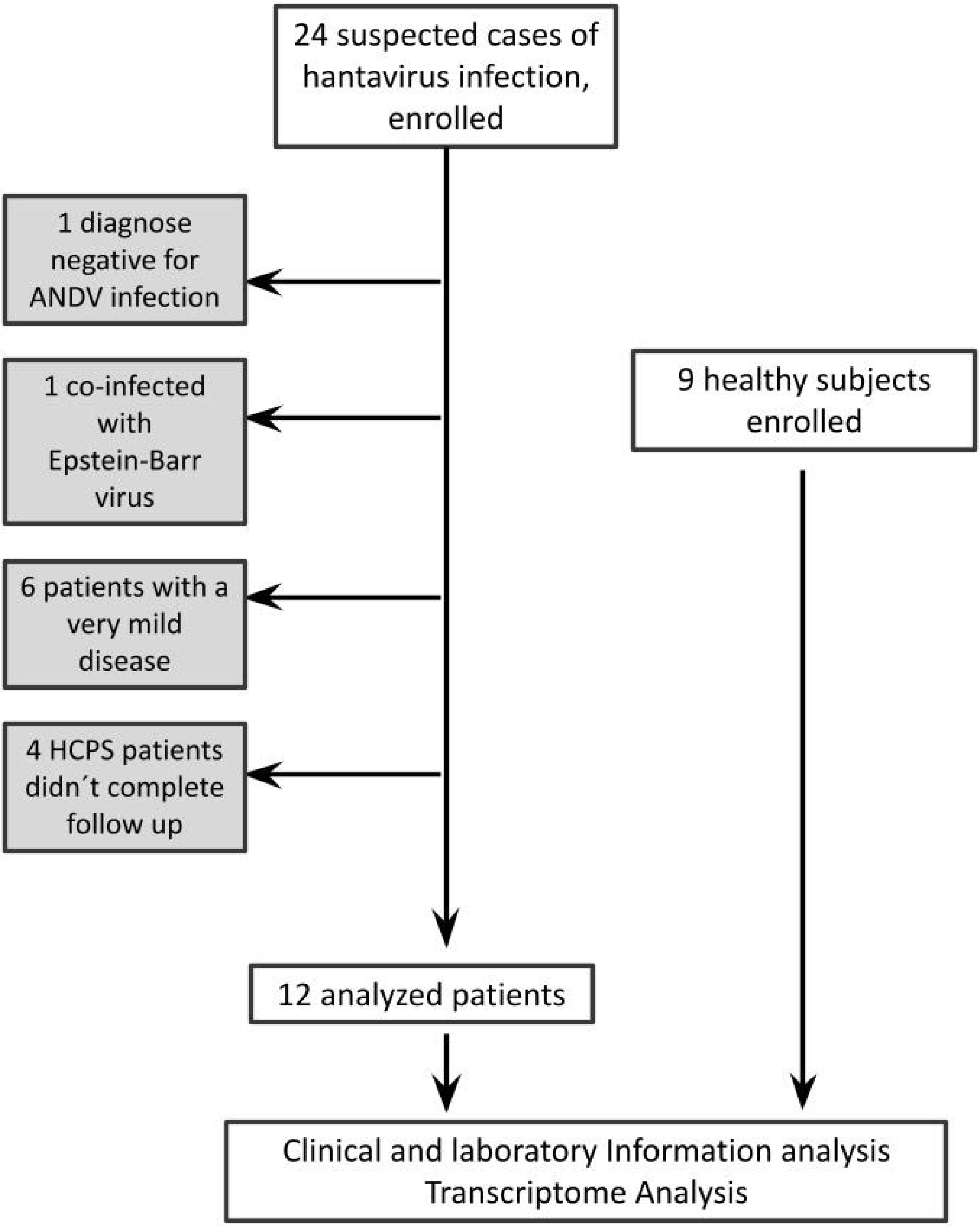


**S1 Fig. Schematic overview of enrolled and analyzed individuals for this study.**

**Supplementary Table 1**. Number of sequenced samples of HCPS patients and Healthy controls.

| | MV+VD (n=5) | ECMO/Fatal (n=7) | Healthy Controls (n=9) |
|---|---|---|---|
| **Early** | 5 | 7 | 9 |
| **Late** | 4 | 3 | 0 |
| **Convalescent** | 4 | 4 | 0 |
| **Total** | 13 | 14 | 9 |

**Supplementary Table 2.** Significant DEGs belonging to marked BTMs from text Figure 3

| | | | MV+VD | | | | | | ECMO/Fatal | | | | | |
|---|---|---|---|---|---|---|---|---|---|---|---|---|---|---|
| | | | Early | | Late | | Convalescent | | Early | | Late | | Convalescent | |
| | | Gene SYMBOL | log Fold Change | adj. P.Val | log Fold Change | adj. P.Val | log Fold Change | adj. P.Val | log Fold Change | adj. P. Val | log Fold Change | adj. P. Val | log Fold Change | adj.P. Val |
| Interferon Response | Antiviral IFN signature LI.M75 | SERPING1 | 3.570 | 0.197 | 2.996 | 1 | 0.307 | 1 | 5.106 | **0.000** | 3.235 | 0.733 | -0.282 | 1 |
| | | RSAD2 | 1.950 | 1 | 1.502 | 1 | 0.311 | 1 | 4.458 | **0.000** | 2.438 | 1 | -0.373 | 1 |
| | | FCER1A | -6.143 | **0.000** | -3.377 | 0.289 | -0.659 | 1 | -4.353 | **0.000** | -6.243 | **0.001** | -0.989 | 1 |
| | | C1QB | 5.868 | **0.000** | 4.496 | **0.035** | 0.735 | 1 | 4.688 | **0.002** | 6.453 | **0.000** | 0.403 | 1 |
| | | IFIT1 | 1.698 | 1 | 1.516 | 1 | 0.156 | 1 | 4.164 | **0.015** | 1.493 | 1 | -0.504 | 1 |
| | | ANXA3 | 5.806 | **0.018** | 6.250 | **0.021** | 0.521 | 1 | 5.169 | 0.052 | 7.201 | **0.002** | 0.916 | 1 |
| | Type I IFN response LI.M127 | USP18 | 2.684 | 1 | 1.451 | 1 | 0.426 | 1 | 4.006 | **0.000** | 1.768 | 1 | 0.134 | 1 |
| | | RSAD2 | 1.950 | 1 | 1.502 | 1 | 0.311 | 1 | 4.458 | **0.000** | 2.438 | 1 | -0.373 | 1 |
| | | IFIT1 | 1.698 | 1 | 1.516 | 1 | 0.156 | 1 | 4.164 | **0.015** | 1.493 | 1 | -0.504 | 1 |
| | Innate antiviral response LI. M150 | RSAD2 | 1.950 | 1 | 1.502 | 1 | 0.311 | 1 | 4.458 | **0.000** | 2.438 | 1 | -0.373 | 1 |
| | | IFIT1 | 1.698 | 1 | 1.516 | 1 | 0.156 | 1 | 4.164 | **0.015** | 1.493 | 1 | -0.504 | 1 |
| | | OAS3 | 1.365 | 1 | 1.076 | 1 | 0.325 | 1 | 3.012 | 0.087 | 1.032 | 1 | -0.194 | 1 |
| | | OASL | 2.579 | 1 | 2.419 | 1 | 0.425 | 1 | 3.273 | 0.097 | 1.898 | 1 | 0.172 | 1 |
| Dendritic cells | Activated dendritic cells LI. M67 | CD38 | 2.692 | 0.297 | 2.238 | 1 | 0.147 | 1 | 3.536 | **0.000** | 2.767 | 0.311 | 0.696 | 1 |
| | | SERPING1 | 3.570 | 0.197 | 2.996 | 1 | 0.307 | 1 | 5.106 | **0.000** | 3.235 | 0.733 | -0.282 | 1 |
| | | RSAD2 | 1.950 | 1 | 1.502 | 1 | 0.311 | 1 | 4.458 | **0.000** | 2.438 | 1 | -0.373 | 1 |
| | | C1QC | 6.608 | **0.000** | 5.569 | **0.013** | 0.794 | 1 | 5.569 | **0.002** | 7.303 | **0.000** | 0.414 | 1 |
| | | C2 | 2.858 | 0.228 | 2.221 | 1 | 0.671 | 1 | 3.220 | **0.010** | 3.064 | 0.117 | 0.336 | 1 |
| | Enriched in actvated Dendritic cells LI.M165 | IFI27 | 9.372 | **0.000** | 7.934 | **0.000** | 0.963 | 1 | 10.403 | **0.000** | 9.180 | **0.000** | 1.020 | 1 |
| | | SERPING1 | 3.570 | 0.197 | 2.996 | 1 | 0.307 | 1 | 5.106 | **0.000** | 3.235 | 0.733 | -0.282 | 1 |
| | | RSAD2 | 1.950 | 1 | 1.502 | 1 | 0.311 | 1 | 4.458 | **0.000** | 2.438 | 1 | -0.373 | 1 |
| | | SIGLEC1 | 3.389 | 0.273 | 2.408 | 1 | 0.899 | 1 | 4.607 | **0.000** | 2.979 | 1 | -0.248 | 1 |
| | | IFIT3 | 2.179 | 1 | 2.093 | 1 | 0.190 | 1 | 4.338 | **0.004** | 2.222 | 1 | -0.572 | 1 |
| | | HESX1 | 1.943 | 1 | 1.298 | 1 | 0.151 | 1 | 3.701 | **0.011** | 1.880 | 1 | -0.440 | 1 |
| | | IFIT1 | 1.698 | 1 | 1.516 | 1 | 0.156 | 1 | 4.164 | **0.015** | 1.493 | 1 | -0.504 | 1 |
| | | TNFAIP6 | 3.388 | 1 | 4.130 | 0.652 | -0.272 | 1 | 4.707 | **0.039** | 4.548 | 0.145 | 0.147 | 1 |
| | Plasma cell suface signature LI.S3 | KCNN3 | 3.264 | 0.602 | 3.074 | 1 | 1.016 | 1 | 4.974 | **0.000** | 5.112 | **0.000** | 0.960 | 1 |
| | | SDC1 | 3.653 | 0.473 | 3.589 | 1 | 0.280 | 1 | 5.454 | **0.000** | 4.965 | **0.010** | -0.045 | 1 |
| | | CAV1 | 2.169 | 1 | 2.174 | 1 | 0.384 | 1 | 3.548 | 0.068 | 4.091 | **0.013** | 0.228 | 1 |

adj.P.Val < 0,05 in **bold**

**Supplementary Table 3.** Gene enrichment analysis: Biological processes and enriched genes in ECMO/Fatal group of patients in early-acute-response, from Figure 4B

| Enriched in ECMO/Fatal early-acute-phase | | | | | | |
|---|---|---|---|---|---|---|
| **ID** | **Description** | **Gene Ratio** | **pval** | **p.adjust** | **qval** | **geneID** |
| GO:0060337 | type I interferon signaling pathway | 8/44 | 5.07E-11 | 1.18E-08 | 8.62E-09 | USP18/MX1/RSAD2/IFI6/IFIT3/ISG15/IFIT1/IFITM3 |
| GO:0071357 | cellular response to type I interferon | 8/44 | 5.07E-11 | 1.18E-08 | 8.62E-09 | USP18/MX1/RSAD2/IFI6/IFIT3/ISG15/IFIT1/IFITM3 |
| GO:0034340 | response to type I interferon | 8/44 | 7.10E-11 | 1.18E-08 | 8.62E-09 | USP18/MX1/RSAD2/IFI6/IFIT3/ISG15/IFIT1/IFITM3 |
| GO:0045071 | negative regulation of viral genome replication | 5/44 | 2.62E-07 | 3.27E-05 | 2.38E-05 | MX1/RSAD2/ISG15/IFIT1/IFITM3 |
| GO:1903901 | negative regulation of viral life cycle | 5/44 | 1.64E-06 | 0.000163 | 0.0001194 | MX1/RSAD2/ISG15/IFIT1/IFITM3 |
| GO:0045069 | regulation of viral genome replication | 5/44 | 2.85E-06 | 0.000236 | 0.0001728 | MX1/RSAD2/ISG15/IFIT1/IFITM3 |
| GO:0048525 | negative regulation of viral process | 5/44 | 3.49E-06 | 0.000248 | 0.0001815 | MX1/RSAD2/ISG15/IFIT1/IFITM3 |

**Supplementary Table 4.** Correlations between transcriptional profile enriched in ECMO/Fatal patients in an early acute response and clinical data.

| Patient data | variable | Spearman r | p value |
|---|---|---|---|
| Severity | LI.M75 | 0.85689307 | 0.00528675 |
| Viral.load | LI.M75 | 0.76190476 | 0.03596737 |
| Platelets | LI.M75 | 0.43432641 | 0.74963232 |
| Neutrophiles | LI.M75 | -0.2505701 | 0.06223806 |
| Total Leuk | LI.M75 | 0.06694619 | 0.99071003 |
| CD3 | LI.M75 | 0.23333333 | 0.22251962 |
| LB_CD19 | LI.M75 | 0.25 | 0.41413315 |
| NK | LI.M75 | 0.5 | 0.02880326 |
| ThL_CD4 | LI.M75 | 0.65 | 0.05350696 |
| LTCD8 | LI.M75 | 0.21666667 | 0.78445385 |
| NK_Bri. | LI.M75 | 0.63333333 | 0.1935741 |
| NKT | LI.M75 | 0.45 | 0.06822605 |
| First Symptoms (Days) | LI.M75 | -0.2576726 | 0.48721103 |
| Days_in_SC | LI.M75 | -0.1773937 | 0.57407531 |
| Days_Hosp | LI.M75 | -0.1225921 | 0.77513642 |
| Days_Oxigen | LI.M75 | 0.20000123 | 0.924808 |
| Days_VD | LI.M75 | 0.16372699 | 0.76477612 |
| Days_VM | LI.M75 | 0.26149879 | 0.78619544 |
| Days_ECMO | LI.M75 | 0.49234529 | 0.50664177 |
| Severity | LI.M127 | 0.70999712 | 0.01931519 |
| Viral.load | LI.M127 | 0.71428571 | 0.01910081 |
| Platelets | LI.M127 | 0.38879219 | 0.77663659 |
| Neutrophiles | LI.M127 | -0.0683373 | 0.02825912 |
| Total Leuk | LI.M127 | 0.15062893 | 0.99875448 |
| CD3 | LI.M127 | 0.13333333 | 0.27587925 |
| LB_CD19 | LI.M127 | 0.11666667 | 0.61444963 |
| NK | LI.M127 | 0.43333333 | 0.043599 |
| ThL_CD4 | LI.M127 | 0.55 | 0.0917131 |
| LTCD8 | LI.M127 | 0.1 | 0.80277039 |
| NK_Bri. | LI.M127 | 0.66666667 | 0.31869205 |
| NKT | LI.M127 | 0.35 | 0.0336826 |
| First Symptoms (Days) | LI.M127 | -0.2254635 | 0.48459695 |
| Days_in_SC | LI.M127 | -0.2956562 | 0.71677523 |
| Days_Hosp | LI.M127 | -0.0875658 | 0.68027089 |
| Days_Oxigen | LI.M127 | 0.1649133 | 0.96623331 |
| Days_VD | LI.M127 | 0.14593058 | 0.70169464 |

| | | | |
|---|---|---|---|
| Days_VM | LI.M127 | 0.26503256 | 0.87579895 |
| Days_ECMO | LI.M127 | 0.5118828 | 0.57998318 |
| Severity | LIM.150 | 0.85689307 | 0.00329104 |
| Viral.load | LIM.150 | 0.76190476 | 0.01970578 |
| Platelets | LIM.150 | 0.43082378 | 0.54171792 |
| Neutrophiles | LIM.150 | -0.2095677 | 0.03529088 |
| Total Leuk | LIM.150 | 0.05020965 | 0.78512406 |
| CD3 | LIM.150 | 0.11666667 | 0.36599842 |
| LB_CD19 | LIM.150 | 0.11666667 | 0.62433739 |
| NK | LIM.150 | 0.36666667 | 0.04832106 |
| ThL_CD4 | LIM.150 | 0.55 | 0.09782672 |
| LTCD8 | LIM.150 | 0.11666667 | 0.98885497 |
| NK_Bri. | LIM.150 | 0.55 | 0.29458124 |
| NKT | LIM.150 | 0.35 | 0.04739689 |
| First Symptoms (Days) | LIM.150 | -0.300618 | 0.50274785 |
| Days_in_SC | LIM.150 | -0.2069593 | 0.79472621 |
| Days_Hosp | LIM.150 | -0.038529 | 0.88558049 |
| Days_Oxigen | LIM.150 | 0.29824745 | 0.90044533 |
| Days_VD | LIM.150 | 0.23491264 | 0.55384287 |
| Days_VM | LIM.150 | 0.36044428 | 0.78353866 |
| Days_ECMO | LIM.150 | 0.53532782 | 0.48073403 |
| Severity | LI.M67 | 0.70999712 | 0.0172295 |
| Viral.load | LI.M67 | 0.76190476 | 0.00747737 |
| Platelets | LI.M67 | 0.27320532 | 0.81557709 |
| Neutrophiles | LI.M67 | -0.2323468 | 0.06489079 |
| Total Leuk | LI.M67 | 0.06694619 | 0.56925328 |
| CD3 | LI.M67 | 0.23333333 | 0.34494149 |
| LB_CD19 | LI.M67 | 0.25 | 0.37027055 |
| NK | LI.M67 | 0.5 | 0.04464802 |
| ThL_CD4 | LI.M67 | 0.65 | 0.04571352 |
| LTCD8 | LI.M67 | 0.21666667 | 0.89107344 |
| NK_Bri. | LI.M67 | 0.63333333 | 0.13118005 |
| NKT | LI.M67 | 0.45 | 0.11053569 |
| First Symptoms (Days) | LI.M67 | -0.3757725 | 0.2468725 |
| Days_in_SC | LI.M67 | -0.2956562 | 0.52503369 |
| Days_Hosp | LI.M67 | -0.1926448 | 0.61190966 |
| Days_Oxigen | LI.M67 | 0.04210552 | 0.7583034 |
| Days_VD | LI.M67 | 0.07118565 | 0.96473865 |
| Days_VM | LI.M67 | 0.1307494 | 0.89280923 |
| Days_ECMO | LI.M67 | 0.38684273 | 0.76018116 |
| Severity | LI.M165 | 0.6610318 | 0.0094674 |
| Viral.load | LI.M165 | 0.47619048 | 0.00473133 |

| | | | |
|---|---|---|---|
| Platelets | LI.M165 | 0.59895013 | 0.40717226 |
| Neutrophiles | LI.M165 | 0.10933969 | 0.18888938 |
| Total Leuk | LI.M165 | -0.3179944 | 0.34193769 |
| CD3 | LI.M165 | -0.3333333 | 0.86327871 |
| LB_CD19 | LI.M165 | -0.3333333 | 0.88737305 |
| NK | LI.M165 | 0.08333333 | 0.20183057 |
| ThL_CD4 | LI.M165 | 0.13333333 | 0.24996697 |
| LTCD8 | LI.M165 | -0.35 | 0.46462202 |
| NK_Bri. | LI.M165 | 0.25 | 0.36285796 |
| NKT | LI.M165 | 0.15 | 0.17909994 |
| First Symptoms (Days) | LI.M165 | -0.3185119 | 0.58628249 |
| Days_in_SC | LI.M165 | -0.0886969 | 0.88023967 |
| Days_Hosp | LI.M165 | 0.03502632 | 0.96299897 |
| Days_Oxigen | LI.M165 | 0.28070348 | 0.75513644 |
| Days_VD | LI.M165 | 0.22779407 | 0.49171035 |
| Days_VM | LI.M165 | 0.303904 | 0.65819659 |
| Days_ECMO | LI.M165 | 0.60957036 | 0.34653176 |
| Severity | LI.S3 | 0.6610318 | 0.00303832 |
| Viral.load | LI.S3 | 0.5 | 0.46641958 |
| Platelets | LI.S3 | -0.0105079 | 0.70273403 |
| Neutrophiles | LI.S3 | 0.10478387 | 0.93656897 |
| Total Leuk | LI.S3 | 0.05857792 | 0.68341263 |
| CD3 | LI.S3 | 0.33333333 | 0.68667859 |
| LB_CD19 | LI.S3 | 0.48333333 | 0.43827637 |
| NK | LI.S3 | 0.61666667 | 0.16362994 |
| ThL_CD4 | LI.S3 | 0.63333333 | 0.24142795 |
| LTCD8 | LI.S3 | 0.18333333 | 0.70582497 |
| NK_Bri. | LI.S3 | 0.85 | 0.20580379 |
| NKT | LI.S3 | 0.45 | 0.62648809 |
| First Symptoms (Days) | LI.S3 | -0.0143151 | 0.62237011 |
| Days_in_SC | LI.S3 | -0.620878 | 0.08455317 |
| Days_Hosp | LI.S3 | -0.1856395 | 0.71076516 |
| Days_Oxigen | LI.S3 | 0.00350879 | 0.43641078 |
| Days_VD | LI.S3 | -0.1423713 | 0.50712977 |
| Days_VM | LI.S3 | 0.12014809 | 0.31381571 |
| Days_ECMO | LI.S3 | 0.3360452 | 0.1875933 |
| Severity | LI.M156.0 | 0.22034393 | 0.34995296 |
| Viral.load | LI.M156.0 | -0.4285714 | 0.1998436 |
| Platelets | LI.M156.0 | -0.0245184 | 0.4789934 |
| Neutrophiles | LI.M156.0 | 0.1548979 | 0.36224943 |
| Total Leuk | LI.M156.0 | -0.2092069 | 0.82345475 |
| CD3 | LI.M156.0 | 0.18333333 | 0.60707643 |

| | | | |
|---|---|---|---|
| LB_CD19 | LI.M156.0 | 0.25 | 0.31733328 |
| NK | LI.M156.0 | 0.38333333 | 0.77684008 |
| ThL_CD4 | LI.M156.0 | 0.33333333 | 0.6718134 |
| LTCD8 | LI.M156.0 | -0.0166667 | 0.67959823 |
| NK_Bri. | LI.M156.0 | 0.35 | 0.44082141 |
| NKT | LI.M156.0 | 0.1 | 0.37780305 |
| First Symptoms (Days) | LI.M156.0 | -0.0644181 | 0.92211177 |
| Days_in_SC | LI.M156.0 | -0.3547874 | 0.20268935 |
| Days_Hosp | LI.M156.0 | 0.28721585 | 0.12109956 |
| Days_Oxigen | LI.M156.0 | 0.03157914 | 0.49104207 |
| Days_VD | LI.M156.0 | 0.20999766 | 0.43674399 |
| Days_VM | LI.M156.0 | 0.06714158 | 0.4352826 |
| Days_ECMO | LI.M156.0 | 0.1680226 | 0.42333503 |
| Severity | ECMO/Fatal | 0.85689307 | 0.00039188 |
| Viral.load | ECMO/Fatal | 0.78571429 | 0.04842799 |
| Platelets | ECMO/Fatal | 0.42381851 | 0.58986031 |
| Neutrophiles | ECMO/Fatal | -0.1138955 | 0.32511055 |
| Total Leuk | ECMO/Fatal | -0.0753145 | 0.42909779 |
| CD3 | ECMO/Fatal | -0.0666667 | 0.70703572 |
| LB_CD19 | ECMO/Fatal | -0.0333333 | 0.59223916 |
| NK | ECMO/Fatal | 0.2 | 0.11415758 |
| ThL_CD4 | ECMO/Fatal | 0.36666667 | 0.16372237 |
| LTCD8 | ECMO/Fatal | -0.0666667 | 0.55513941 |
| NK_Bri. | ECMO/Fatal | 0.45 | 0.22584841 |
| NKT | ECMO/Fatal | 0.18333333 | 0.2492875 |
| First Symptoms (Days) | ECMO/Fatal | -0.2147271 | 0.72218041 |
| Days_in_SC | ECMO/Fatal | -0.2660906 | 0.26074672 |
| Days_Hosp | ECMO/Fatal | -0.0945711 | 0.96723995 |
| Days_Oxigen | ECMO/Fatal | 0.23158037 | 0.72085776 |
| Days_VD | ECMO/Fatal | 0.21355694 | 0.59707709 |
| Days_VM | ECMO/Fatal | 0.30743777 | 0.57542645 |
| Days_ECMO | ECMO/Fatal | 0.54705032 | 0.30720828 |
| Severity | VM+VD | -0.6610318 | 0.01928447 |
| Viral.load | VM+VD | -0.4761905 | 0.07088801 |
| Platelets | VM+VD | -0.5253949 | 0.74920103 |
| Neutrophiles | VM+VD | -0.127563 | 0.54213733 |
| Total Leuk | VM+VD | 0.2259434 | 0.40337764 |
| CD3 | VM+VD | 0.01666667 | 0.513567 |
| LB_CD19 | VM+VD | 0 | 0.35053468 |
| NK | VM+VD | -0.3666667 | 0.14110461 |
| ThL_CD4 | VM+VD | -0.4333333 | 0.10349041 |
| LTCD8 | VM+VD | 0.13333333 | 0.74048611 |

| | | | |
|---|---|---|---|
| NK_Bri. | VM+VD | -0.5166667 | 0.10781202 |
| NKT | VM+VD | -0.4166667 | 0.40738376 |
| First Symptoms (Days) | VM+VD | 0.50460875 | 0.35567564 |
| Days_in_SC | VM+VD | 0.11826248 | 0.59460916 |
| Days_Hosp | VM+VD | -0.1225921 | 0.96084685 |
| Days_Oxigen | VM+VD | -0.4491256 | 0.4579917 |
| Days_VD | VM+VD | -0.1281342 | 0.63456716 |
| Days_VM | VM+VD | -0.4063833 | 0.45495355 |
| Days_ECMO | VM+VD | -0.5353278 | 0.29475128 |
| Severity | | | |

**Supplementary Table 5.** Cell surface markers used to define WBC populations by flow cytometry immunophenotyping.

| **Subpopulation** | **Surface Markers** |
|---|---|
| CD8 T cells | CD45+ CD3+ CD8+ |
| CD4 T cells | CD45+ CD3+ CD4+ |
| B cells | CD45+ CD3- CD19+ |
| NK cells | CD45+ CD3- CD56+ |
| NKT cells | CD45+ CD3+ CD56+ |
